# Incidence, risk factors, and clinical outcomes of mannitol-induced acute kidney injury among acute stroke patients admitted to East Avenue Medical Center

**DOI:** 10.1101/2023.07.02.23292143

**Authors:** Ralph Jefferson T. Ramos, Karen S. Cabigas, Elyzel C. Rabara, Marissa Elizabeth Lim

## Abstract

**Background:** In the Philippines, stroke is the second most common cause of mortality and is among the five leading causes of disability. Mannitol, a hyperosmolar agent, is a mainstay for the treatment of brain edema caused by elevated intracranial pressure in acute stroke patients. It is a potent diuretic that may trigger intravascular volume depletion, electrolyte imbalance, and renal tubular damage, which may lead to acute kidney injury in acute stroke.

**Objective:** This study aims to describe the incidence, identify the risk factors, and determine the clinical outcomes of mannitol-induced acute kidney injury among acute stroke patients admitted to East Avenue Medical Center from January 1, 2019, to December 31, 2021.

**Methods:** This is a retrospective study conducted at the East Avenue Medical Center, a tertiary training hospital in Quezon City, Philippines. A three-year chart review of acute stroke patients who developed mannitol-induced acute kidney injury (MI-AKI) was conducted.

**Result:** A total of 348 eligible acute stroke patients were included in the study. Of these, 60 patients (17%) developed MI-AKI during confinement. There was a higher predominance among males than females with more than half of patients (65%) belonging to the 40 to 59 years age group. The risk factors identified were high National Institutes of Health Stroke Scale (NIHSS) score, chronic kidney disease, possibly cardiovascular disease, such as heart failure, intraparenchymal type hemorrhagic stroke, and high mannitol infusion dose >0.40g/kg in more than 48 hours. A high mortality rate was observed among MI-AKI patients in the study as compared with those without AKI (61.67% vs 27.43%, P<0.001) whereas spontaneous resolution of AKI was seen in 25 patients (41.67%, P<0.001). Only 4 patients (6.67%, P=0.001) who had renal replacement therapy had worsening renal function compared to the 27 patients (45%, P<0.001) who did not receive renal replacement therapy.

**Conclusion:** Risk factors for MI-AKI include moderate to severe NIHSS score, chronic kidney disease 3 and above, cardiovascular disease, and high-dose mannitol infusion. Early nephrology referral among patients with mannitol infusion for stroke is a must among patients who have co-morbidities including chronic kidney disease stage 3 and above, and cardiovascular disease such as heart failure. Serial monitoring of renal function should be performed especially for those patients with moderate to severe NIHSS scores and high-dose mannitol infusion for appropriate dosing to prevent MI-AKI.

## Introduction

Stroke remains the Philippines’ second greatest cause of mortality and was among the top five causes of disability from 2009 to 2019. Stroke prevalence estimates range from 0.9% (2005) to 2.6% (2017) of the general population. While seven out of ten strokes are classified as ischemic, the remaining three are classified as hemorrhagic (1). Among Filipino stroke patients undergoing thrombolysis, in-hospital mortality has been reported to be 14.6%, and in those who have been discharged, death occurred in 3.7% (2). As a result of variability in data collection and inadequate epidemiological studies, there is currently limited data on the etiological factors that lead to death among Filipino stroke patients.

In a study conducted by Shin-Yi Lin et al 2015, a major cause of long-term morbidity and mortality among acute stroke patients is the development of cerebral edema. Mannitol, a hyperosmolar diuretic, has been established as a treatment for acute cerebral edema (3). It is a potent diuretic that decreases elevated intracranial pressure in patients with brain edema by promoting the osmotic flow of water out of the cerebral parenchyma and by autoregulatory vasoconstriction. It also augments cerebral blood flow (4). Although beneficial in many cases, mannitol is known to alter intravascular volume status, serum osmolarity, electrolytes, creatinine, and blood urea nitrogen and increase the risk of acute kidney injury (AKI) (4). Especially in high doses (>0.20 kg/day or cumulative dose of >0.40 kg in 48 hours), mannitol has been found to cause acute tubular necrosis when serum osmolarity is above 320 Osm (4).

Mannitol-induced AKI (MI-AKI), also known as mannitol-induced osmotic nephrosis, is characterized by structural changes that occur at a cellular level in the kidney, primarily at the proximal tubule. The alterations include intracytoplasmic vacuolization and distension of tubular cells (5). The form of acute renal failure in MI-AKI is often observed to be oliguric/anuric and may be asymptomatic or may mimic other nephrotic conditions (5). The risk factors that have been associated with MI-AKI in stroke patients include diabetes mellitus, underlying renal impairment, high initial NIHSS scores, and concurrent use of diuretics (3). In patients with intracranial hemorrhage, MI-AKI occurred more frequently in those who received mannitol infusion at a rate of ≥ 1.34 g/kg/day compared with patients who received slower rates. In this subset of patients, the independent risk factors for MI-AKI were infusion at a rate of ≥1.34g/kg/day, diastolic blood pressure (DBP) ≥ 100 mmHg, and glomerular filtration rate (GFR) <60ml/min/1.73m^2^ (6).

No local data has been published establishing the burden, causes, and optimal management of MI-AKI among Filipino stroke patients. This study aims to describe the incidence, enumerate the risk factors, and establish the clinical outcome of MI-AKI among acute stroke patients admitted to East Avenue Medical Center (EAMC) from January 1, 2019, to December 31, 2021. Outlining these risk factors may aid in predicting the occurrence of MI-AKI injury among adult acute stroke patients and allow clinicians to create an appropriate plan of management among patients who have the highest risk. Secondarily, we aimed to describe the population of acute stroke patients with MI-AKI in terms of:

- Demographics: age, gender, co-morbidities, medications, and estimated glomerular filtration rate (eGFR) of acute stroke patients who are likely to develop mannitol-induced AKI
- Identify the risk factors for MI-AKI among acute stroke patients treated with mannitol
- Dosage, cumulative dose, and duration of mannitol use among acute stroke patients who developed MI-AKI
- Clinical outcomes such as the resolution of MI-AKI, death, or acute kidney injury requiring renal replacement therapy upon discharge among acute stroke patients with MI-AKI admitted to the EAMC Stroke Unit

## Materials and methods

### Study design

This is a three-year retrospective analysis of hospital medical records dated January 1, 2019, to December 31, at the Section of Nephrology in EAMC 2021. Individual information of the patients was anonymized using code numbers.

### Sampling and sample size

Patients who were 19 years old and above, had either an acute ischemic or hemorrhagic stroke, and developed AKI following mannitol administration beyond the first 48 hours of confinement comprise the study group. Baseline NIHSS scores upon admission and systolic blood pressures were obtained. Patients who were under renal replacement therapy (RRT), had nosocomial infections, had brain tumors, and expired within 48 hours of admission were excluded. After, using G*Power 3.1.9.2, a minimum of 63 patients are required for this study based on a 3.62 odds ratio of diabetic patients who develop MI-AKI (3), with a 5% level of significance and 95% power.

### Data gathering

The data extracted included the hospital codes assigned to the patients with their corresponding age, sex, comorbidities, initial NIHSS scores, medications, GFR as defined by the 2021 Chronic Kidney Disease Epidemiology Collaboration (CKD-EPI) criteria, and mannitol treatment upon admission. A list of medications such as anti-hypertensive medications, nephrotoxic agents, and diuretics was listed. Lastly, the cumulative dose and the duration of mannitol therapy were documented. Clinical outcomes such as the resolution of MI-AKI, unimprovement either with the disposition of AKI-requiring RRT or AKI-not requiring RRT, and death were likewise recorded.

### Data processing and statistical analysis

The collected data were tabulated, encoded, and analyzed using the Stata 13.1 program. Descriptive statistics were used to summarize the demographic and clinical characteristics of the patients. Frequency and proportion were used for categorical variables and mean and standard deviation (SD) for normally distributed continuous variables. Independent sample T-test and Fisher’s exact/Chi-square test were used to determine the difference in mean and frequency, respectively, between patients with and without MI-AKI. The odds ratio and corresponding 95% confidence intervals from binary logistic regression were computed to determine significant predictors for Mannitol-Induced Acute Kidney Injury. All statistical tests were two-tailed tests. Shapiro-Wilk test was used to test the normality of the continuous variables. Missing values were neither replaced nor estimated. Null hypotheses were rejected at a 0.05 level of significance.

**Figure 1.**
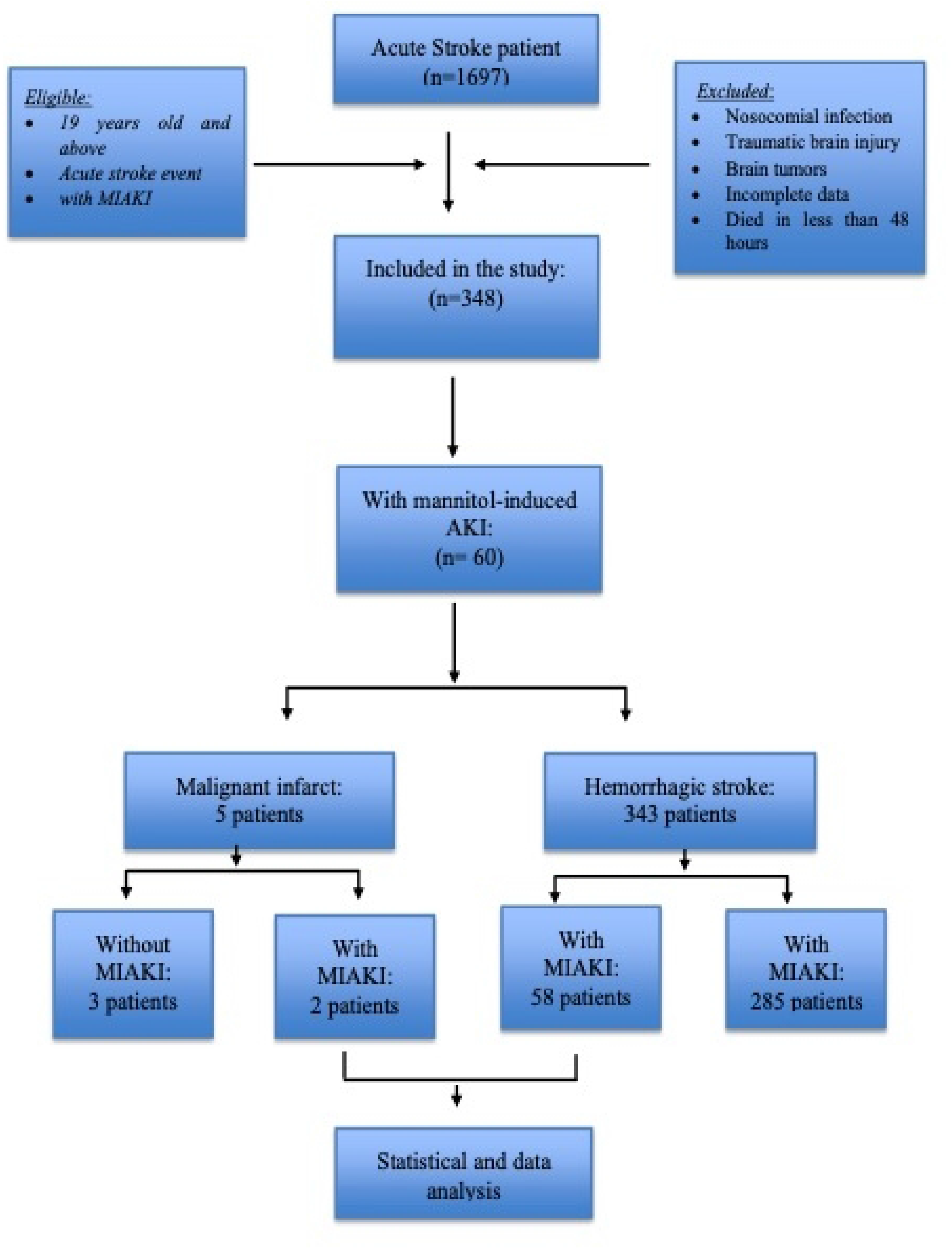
Patient selection process.

## Results

A total of 1,697 patients diagnosed with acute stroke were reviewed, of which 348 (20.5%) patients received mannitol therapy during admission. Most patients had intraparenchymal lesions, 91.09%, followed by subarachnoid hemorrhage, 8.9%, and malignant infarct, 1.44% (Table 1). From the total population, 17% of patients developed MI-AKI (95% confidence interval [CI], 13.42–21.63). MI-AKI occurred after the first 48 hours but not more than 7 days upon administration of mannitol for most patients.

**Table 1.**
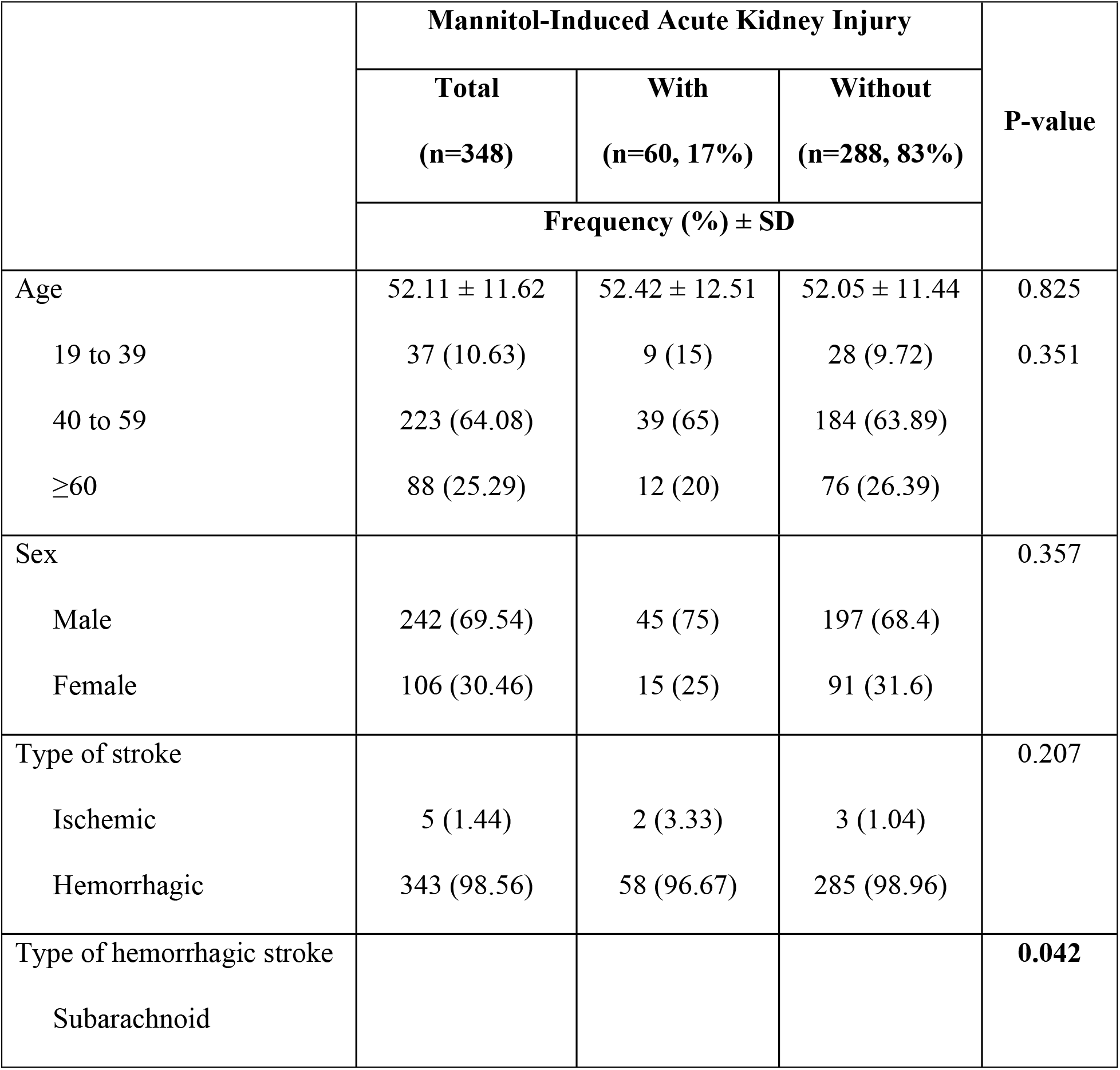
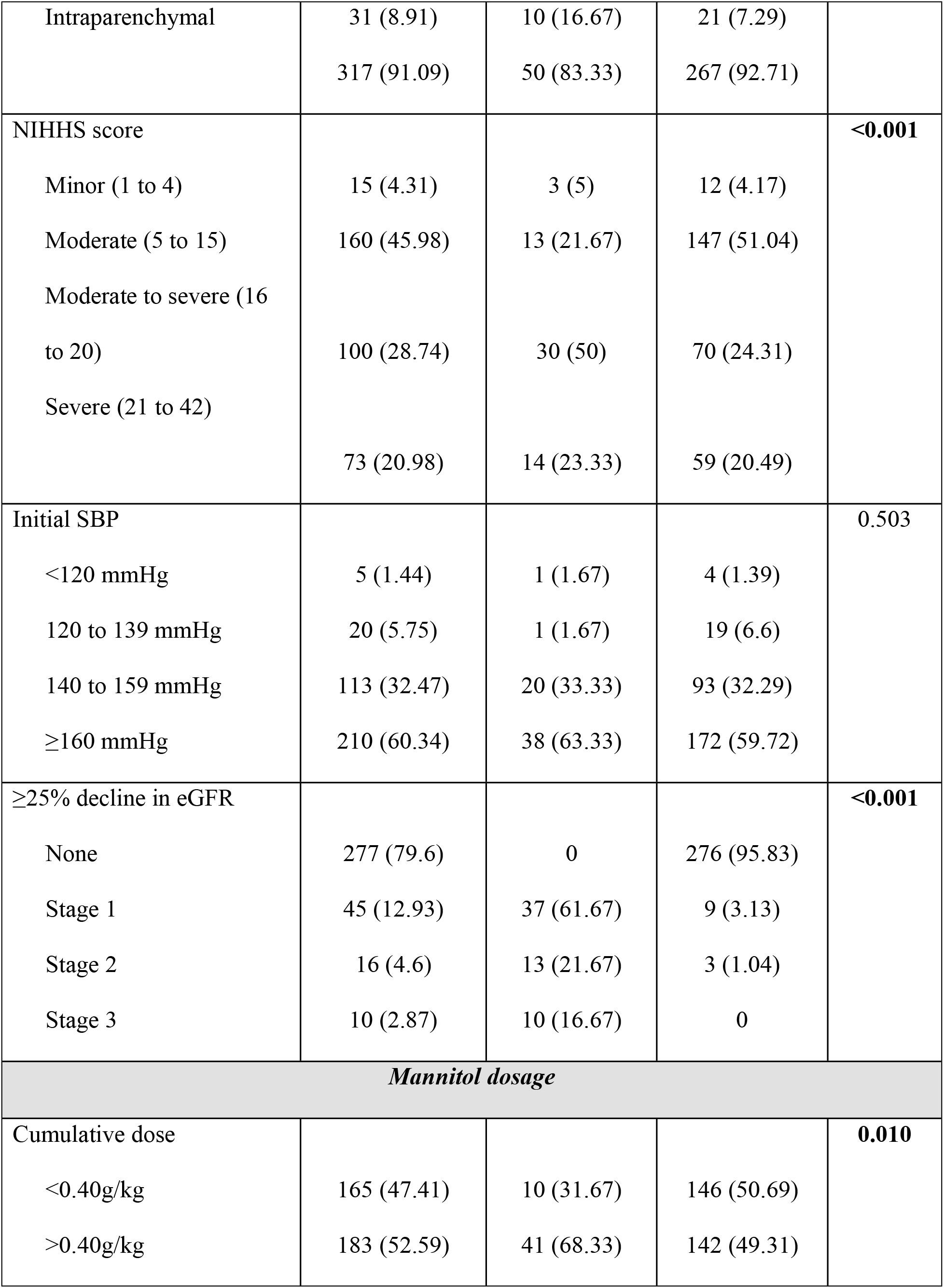
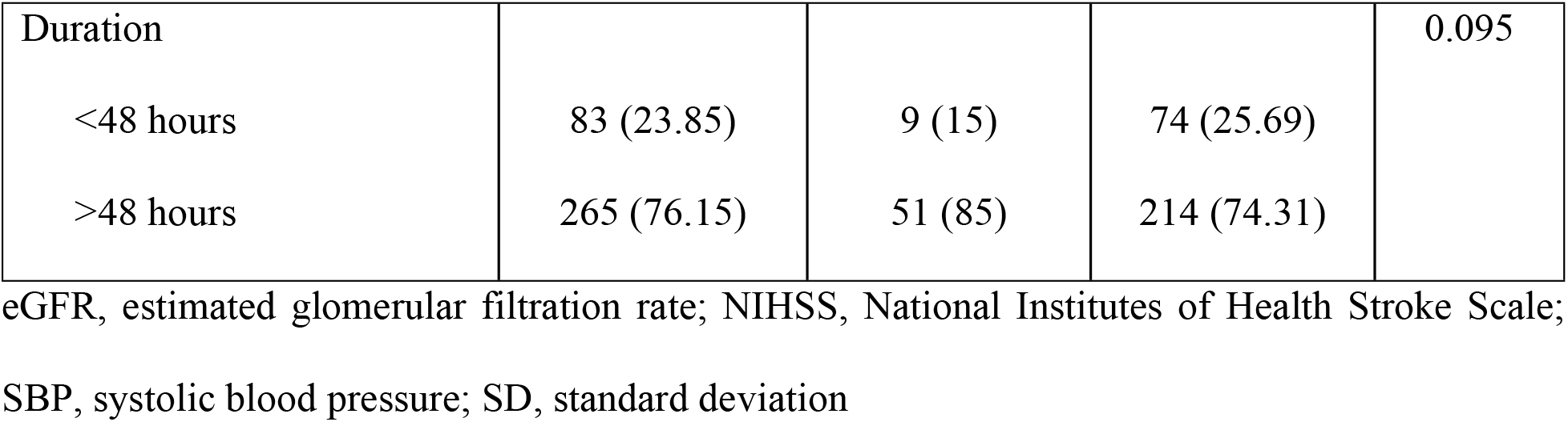
Characteristics of stroke patients who were prescribed mannitol.

|  | Mannitol-Induced Acute Kidney Injury |  |  | P-value |
| --- | --- | --- | --- | --- |
|  | Total<br>(n=348) | With<br>(n=60, 17%) | Without<br>(n=288, 83%) |  |
|  | Frequency (%) ± SD |  |  |  |
| Age | 52.11 ± 11.62 | 52.42 ± 12.51 | 52.05 ± 11.44 | 0.825 |
| 19 to 39 | 37 (10.63) | 9 (15) | 28 (9.72) | 0.351 |
| 40 to 59 | 223 (64.08) | 39 (65) | 184 (63.89) |  |
| ≥60 | 88 (25.29) | 12 (20) | 76 (26.39) |  |
| Sex |  |  |  | 0.357 |
| Male | 242 (69.54) | 45 (75) | 197 (68.4) |  |
| Female | 106 (30.46) | 15 (25) | 91 (31.6) |  |
| Type of stroke |  |  |  | 0.207 |
| Ischemic | 5 (1.44) | 2 (3.33) | 3 (1.04) |  |
| Hemorrhagic | 343 (98.56) | 58 (96.67) | 285 (98.96) |  |
| Type of hemorrhagic stroke |  |  |  | **0.042** |
| Subarachnoid |  |  |  |  |
| Intraparenchymal | 31 (8.91)<br>317 (91.09) | 10 (16.67)<br>50 (83.33) | 21 (7.29)<br>267 (92.71) |  |
| NIHHS score |  |  |  | **<0.001** |
| Minor (1 to 4) | 15 (4.31) | 3 (5) | 12 (4.17) |  |
| Moderate (5 to 15) | 160 (45.98) | 13 (21.67) | 147 (51.04) |  |
| Moderate to severe (16 to 20) | 100 (28.74) | 30 (50) | 70 (24.31) |  |
| Severe (21 to 42) | 73 (20.98) | 14 (23.33) | 59 (20.49) |  |
| Initial SBP |  |  |  | 0.503 |
| <120 mmHg | 5 (1.44) | 1 (1.67) | 4 (1.39) |  |
| 120 to 139 mmHg | 20 (5.75) | 1 (1.67) | 19 (6.6) |  |
| 140 to 159 mmHg | 113 (32.47) | 20 (33.33) | 93 (32.29) |  |
| ≥160 mmHg | 210 (60.34) | 38 (63.33) | 172 (59.72) |  |
| ≥25% decline in eGFR |  |  |  | **<0.001** |
| None | 277 (79.6) | 0 | 276 (95.83) |  |
| Stage 1 | 45 (12.93) | 37 (61.67) | 9 (3.13) |  |
| Stage 2 | 16 (4.6) | 13 (21.67) | 3 (1.04) |  |
| Stage 3 | 10 (2.87) | 10 (16.67) | 0 |  |
| **Mannitol dosage** |  |  |  |  |
| Cumulative dose |  |  |  | **0.010** |
| <0.40g/kg | 165 (47.41) | 10 (31.67) | 146 (50.69) |  |
| >0.40g/kg | 183 (52.59) | 41 (68.33) | 142 (49.31) |  |
| Duration |  |  |  | 0.095 |
| <48 hours | 83 (23.85) | 9 (15) | 74 (25.69) |  |
| >48 hours | 265 (76.15) | 51 (85) | 214 (74.31) |  |

eGFR, estimated glomerular filtration rate; NIHSS, National Institutes of Health Stroke Scale;
SBP, systolic blood pressure; SD, standard deviation

Patients who developed MI-AKI were mostly male (75%) and were 40 to 59 years old (65%). Most patients with MI-AKI (96.67%) were diagnosed with acute hemorrhagic stroke, and of these, a majority (83.33%) were intraparenchymal (P=0.042). About 63% of patients who developed MI-AKI had an initial systolic blood pressure of ≥160 mmHg while 50% had moderate to severe NIHHS scores (P=<0.001). During mannitol treatment, 61.67% developed stage 1 AKI (P≤0.001). About 68% with AKI had a cumulative dose of mannitol >0.40 g/kg (P=0.010) and 51 (85%) patients received mannitol for >48 hours (P=0.095).

Univariate analysis showed that cardiovascular disease (P=0.029) and chronic kidney disease (CKD, P<0.001) were significantly associated with MI-AKI (Table 2A). A subgroup analysis of patients with CKD revealed that stages 3a to 4 had an increased incidence of AKI compared with stages 1 and 2, while stage 5 had no significant association most likely due to the early discontinuation of mannitol upon disease recognition (Table 2A). Among the medications evaluated, anti-hypertensives and diuretics did not demonstrate a significant association with MI-AKI (Table 2B).

**Table 2A.**
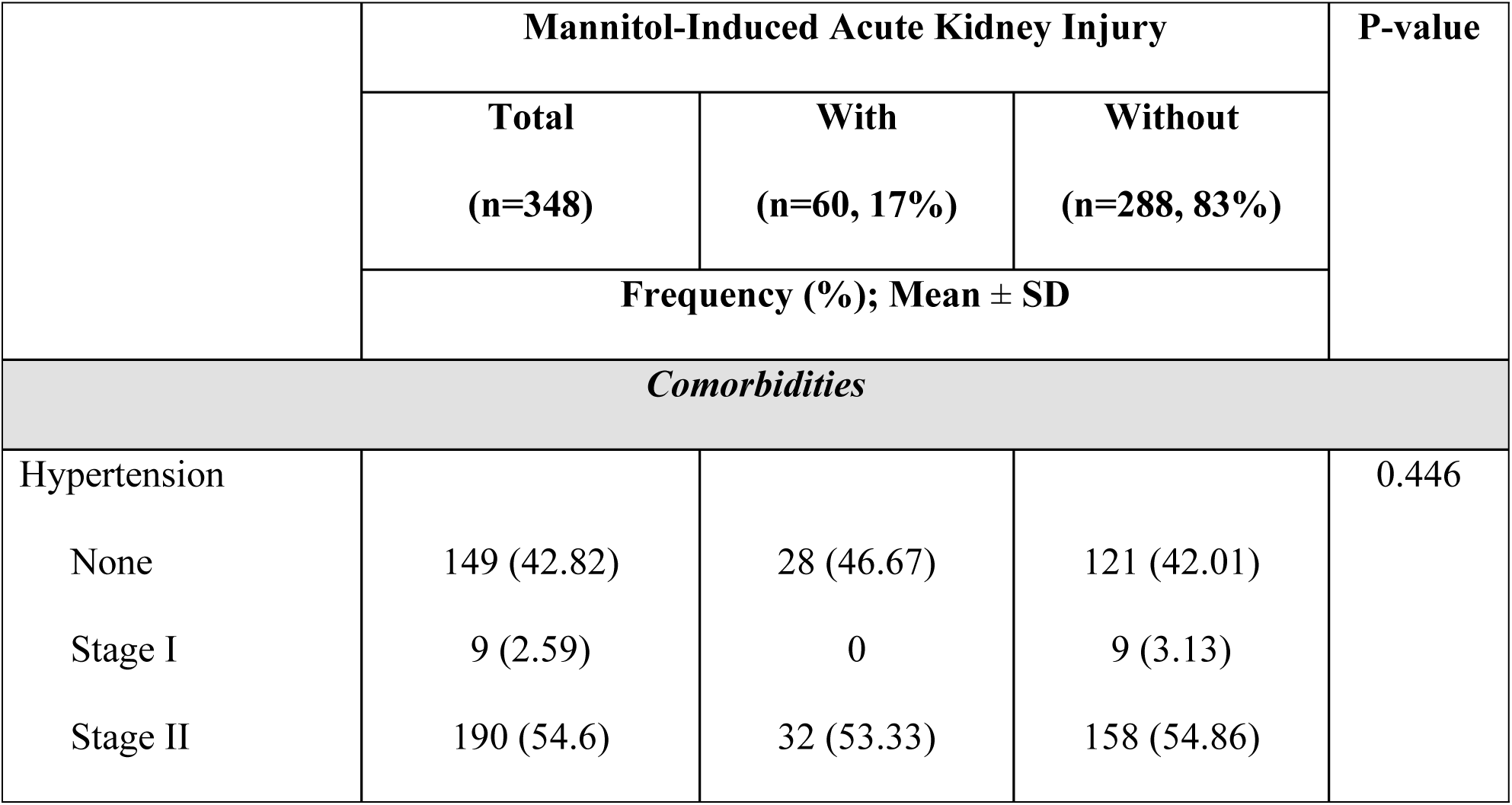
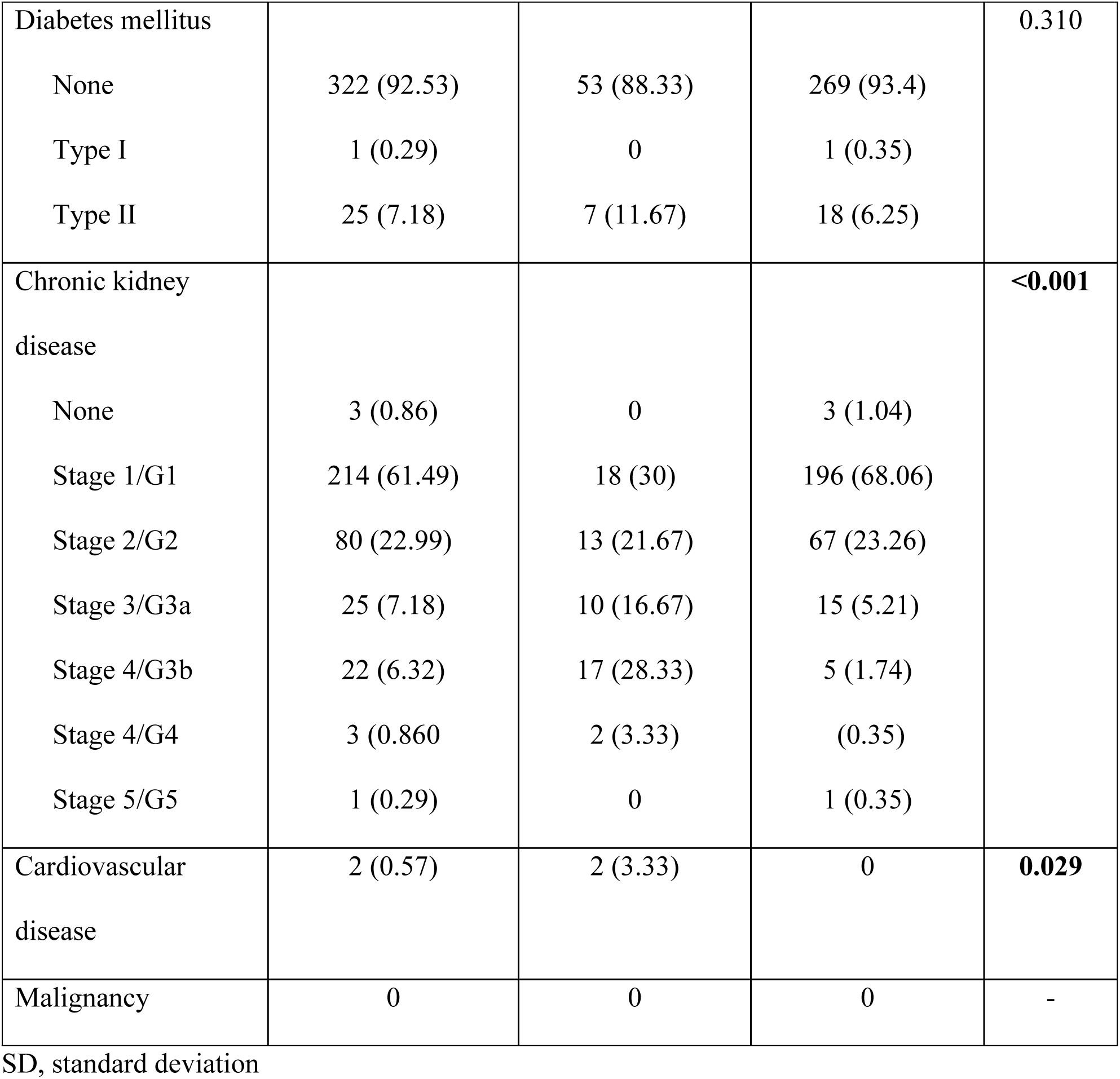
Co-morbidities.

|  | Mannitol-Induced Acute Kidney Injury |  |  | P-value |
| --- | --- | --- | --- | --- |
|  | Total<br>(n=348) | With<br>(n=60, 17%) | Without<br>(n=288, 83%) |  |
|  | Frequency (%); Mean ± SD |  |  |  |
| Comorbidities |  |  |  |  |
| Hypertension |  |  |  | 0.446 |
| None | 149 (42.82) | 28 (46.67) | 121 (42.01) |  |
| Stage I | 9 (2.59) | 0 | 9 (3.13) |  |
| Stage II | 190 (54.6) | 32 (53.33) | 158 (54.86) |  |
| Diabetes mellitus |  |  |  | 0.310 |
| None | 322 (92.53) | 53 (88.33) | 269 (93.4) |  |
| Type I | 1 (0.29) | 0 | 1 (0.35) |  |
| Type II | 25 (7.18) | 7 (11.67) | 18 (6.25) |  |
| Chronic kidney disease |  |  |  | **<0.001** |
| None | 3 (0.86) | 0 | 3 (1.04) |  |
| Stage 1/G1 | 214 (61.49) | 18 (30) | 196 (68.06) |  |
| Stage 2/G2 | 80 (22.99) | 13 (21.67) | 67 (23.26) |  |
| Stage 3/G3a | 25 (7.18) | 10 (16.67) | 15 (5.21) |  |
| Stage 4/G3b | 22 (6.32) | 17 (28.33) | 5 (1.74) |  |
| Stage 4/G4 | 3 (0.86) | 2 (3.33) | (0.35) |  |
| Stage 5/G5 | 1 (0.29) | 0 | 1 (0.35) |  |
| Cardiovascular disease | 2 (0.57) | 2 (3.33) | 0 | **0.029** |
| Malignancy | 0 | 0 | 0 | - |

SD, standard deviation

**Table 2B.** Medications.

|  | Mannitol-Induced Acute Kidney Injury |  |  | P-value |
| --- | --- | --- | --- | --- |
|  | Total<br>(n=348) | With<br>(n=60, 17%) | Without<br>(n=288, 83%) |  |
|  | Frequency (%); Mean ± SD |  |  |  |
| Medication |  |  |  |  |
| Anti-hypertensive | 3 (0.86) | 1 (1.67) | 2 (0.69) | 0.434 |
| Diuretics | 2 (0.57) | 1 (1.67) | 1 (0.35) | 0.316 |
| Nephrotoxic agents | 0 | 0 | 0 | - |

SD, standard deviation

Regarding final patient outcomes, about 62% of patients who had AKI died while 38.33% recovered from acute stroke (P<0.001). Spontaneous recovery with a return to baseline eGFR was observed in 41.67% of patients (P<0.001). On the other hand, worsening of renal function without RRT was observed in 45% of patients (P≤0.001) while 6.67% of patients underwent RRT (P=0.001) (Table 3).

**Table 3.** The outcome of patients with MI-AKI.

|  | Mannitol-Induced Acute Kidney Injury |  |  | P-value |
| --- | --- | --- | --- | --- |
|  | Total<br>(n=348) | With<br>(n=60, 17%) | Without<br>(n=288, 83%) |  |
|  | Frequency (%) |  |  |  |
| Mortality |  |  |  | <0.001 |
| Alive | 232 (66.67) | 23 (38.33) | 209 (72.57) |  |
| Expired | 116 (33.33) | 37 (61.67) | 79 (27.43) |  |
| AKI | 67 (19.25) | 59 (98.33) | 8 (2.78) | **<0.001** |
| Resolution of AKI | 32 (9.20) | 25 (41.67) | 7 (2.43) | **<0.001** |
| Worsening of renal function without RRT | 32 (9.20) | 27 (45.00) | 5 (1.74) | **<0.001** |
| Worsening of renal function with RRT | 4 (1.15) | 4 (6.67) | 0 | **0.001** |

AKI, acute kidney injury; RRT, renal replacement therapy

Initial high NIHHS score of moderate to severe was seen to be associated with a higher risk of developing MI-AKI fivefold compared with a moderate NIHHS score (95% CI, 2.3–9.8; P≤0.001) (Table 4). For every CKD stage increase, the odds of having MI-AKI increase by about threefold (95% CI, 2.05–3.6; P≤0.001). Subarachnoid type of acute hemorrhagic stroke was 2.5 times more likely to develop MI-AKI as compared with the intraparenchymal type of hemorrhagic stroke (95% CI, 1.12–5.72; P=0.024). Lastly, a mannitol dose >0.40g/kg cumulative dose for more than 48 hours increased the risk of having MI-AKI by twofold as compared with patients with a cumulative dose of <0.40g/kg (95% CI, 1.22–4.00; P=0.008).

**Table 4.** Factors associated with MI-AKI.

| Parameters | Crude odds ratio | 95% CI | P-value |
| --- | --- | --- | --- |
| NIHHS score |  |  |  |
| Minor (1 to 4) | 2.8269 | 0.7066 to 11.309 | 0.142 |
| Moderate (5 to 15) | (reference) | - | - |
| Moderate to severe (16 to 20) | 4.8462 | 2.3818 to 9.8602 | **<0.001** |
| Severe (21 to 42) | 2.6832 | 1.1899 to 6.0503 | **0.017** |
| Chronic kidney disease | 2.7406 | 2.0596 to 3.6468 | **<0.001** |
| Type of hemorrhagic stroke |  |  |  |
| Subarachnoid | 2.5429 | 1.1297 to 5.7238 | **0.024** |
| Intraparenchymal | (reference) | - | - |
| Cumulative dose for 48 hours |  |  |  |
| < 0.40g/kg | (reference) | - | - |
| > 0.40g/kg | 2.2187 | 1.2287 to 4.0063 | **0.008** |

## Discussion

This pilot study of patients admitted for stroke in EAMC outlined the risk factors and outcomes in those who developed MI-AKI. The results showed the following important findings in our patient subjects: firstly, our 17% incidence of MI-AKI appears to be higher compared with the previous studies conducted by Kim et al (10.5%) and Lin et al (6.5%). Secondly, our study supports the current observation that moderate to severe initial NIHSS scores, CKD stage 3a and above, cardiovascular diseases such as heart failure, and high doses of mannitol infusion for prolonged periods are likely independent risk factors for MI-AKI.

Previous studies have described a higher incidence of hemorrhagic stroke among patients who develop MI-AKI (6, 7), while others suggest a link between ischemic stroke and MA-AKI (8). However, the association between the type of stroke, either hemorrhagic or ischemic stroke, there appears to be no association between the type of stroke and the risk of developing MI-AKI (3) as reflected in this study. This indicates that MI-AKI is not an uncommon complication in patients treated for stroke regardless of type.

Only patients treated with mannitol were included in the study, and to facilitate ruling out other causes of AKI in this study, we excluded patients being treated with antibiotics for nosocomial infections, including pneumonia, and pulmonary tuberculosis, as well as patients on renal replacement therapy.

A high baseline NIHSS score has been shown in previous studies to increase the risk for MI-AKI (3). Indeed, in our patients with moderate to severe NIHSS scores recruited in this study, there is an apparent fivefold increase in the risk for MI-AKI. Although the NIHSS is established especially as an instrument to evaluate the severity and prognosis of stroke, the strong association between high NIHSS scores and AKI appears to be a result of the presence of severe illness and concomitant complications (9); hence, our study shares consistent findings reported in the literature. Unfortunately, among the study subjects with severe NIHSS scores, it was a challenge to monitor kidney function throughout patient confinement due to their early demise from severe stroke events. Also, most of the patients who received high-dose mannitol infusion developed MI-AKI regardless of the presence of co-morbidities and high NIHSS scores.

Lin et al and Nomani et al found that with every increase in the stage of CKD, the odds of having MI-AKI increase by 2.7 times compared to patients with normal renal function (3, 5) – a finding that is likewise observed in our population. In the setting of renal impairment, the half-life of mannitol has been seen to extend from 70 to 100 minutes to up to >36 hours causing mannitol to accumulate in the kidneys (3). Mannitol is known to trigger water and reabsorption of salt along the entire renal tubule resulting in electrolyte imbalance and volume depletion further increasing the risk for renal injury in advanced CKD (3). Therefore, our results reflect comparable risk rates with other studies owing to these recognized pathophysiologic mechanisms.

Unlike in previous studies, however, comorbidities including diabetes, hypertension, and the use of anti-hypertensive medications and diuretics were not demonstrated to increase the risk of MI-AKI (3, 5). The use of diuretics (i.e., mannitol) and contrast media in cardiovascular diseases, such as heart failure and acute coronary events, increase the risk for AKI. Contrast-induced AKI is a common complication post percutaneous coronary intervention as a result of contrast nephrotoxicity (10). Mannitol generally brings about excessive intravascular volume expansion, especially when administered in high doses and at rapid rates. In addition, volume depletion may lead to renal vasoconstriction and ultimately acute renal injury (11). Hence, patients being treated for cardiovascular diseases appear to be predictably at risk for MI-AKI as demonstrated in our study. However, because of the small population where we identified only two patients with cardiovascular disease, we are unable to draw a conclusive association. We propose further analyses conducted in larger populations to provide a definitive association between these comorbidities and MI-AKI (9).

The usual dose of mannitol for reduction of intracranial pressure is 0.12 to 0.20 g/kg/day administered intravenously as a 20% solution over 30 to 60 minutes (5). The development of MI-AKI was found to be dose-dependent and may occur in high-dose mannitol infusion (>0.20 g/kg/day or >0.40 g/kg in 48 hours) (5). Thus, monitoring the cumulative dose of mannitol appears to have a notable clinical utility in stroke patients on mannitol. When the mannitol is discontinued upon recognition of AKI, MI-AKI after stroke may be transient. However, some patients may have worsening AKI requiring dialysis. In this study, 41.6% had spontaneous resolution of acute kidney injury, and less than half had worsening renal function without RRT. Only 6.67 % of patients with worsening renal function underwent renal RRT. In patients who develop MI-AKI, discontinuing mannitol and starting dialysis, if necessary, are the clear management options (3).

Lastly, it was found that the mortality rate of patients who developed MI-AKI was high at 61.67%. However, the cause of death and its relation to mannitol-induced AKI were not clearly defined in our subjects. We attribute this high mortality rate to brain herniation syndrome, multiple organ dysfunction, and cardiac complications.

We acknowledge that we mainly identified the risk factors and established the clinical outcomes of MI-AKI among a limited number of patients. The study excluded patients with other medical conditions who developed AKI due to other causes. Further studies may employ serial monitoring of renal function, mandatory diagnostic workup, such as serum osmolarity by spectrometry to calculate the osmolar gap and adequate monitoring of water balance such as fluid input and urine output.

## Conclusion and recommendations

This study showed that MI-AKI is associated with patient-specific factors which include a high initial NIHSS score, chronic kidney disease, and a high dose of mannitol. Cardiovascular disease also appears as a significant risk factor; however, MI-AKI in heart disease is likely due to treatment-associated nephrotoxicity.

Early referral to a renal specialist is recommended for comorbidities including chronic kidney disease stage 3 and above. Serial monitoring of patient renal function should be performed especially for those with initial moderate to severe NIHSS scores and high-dose mannitol infusion. In addition, it is recommended that a multi-center study be done to create a risk stratification tool for the early identification of at-risk patients for MI-AKI.

### Ethical Considerations

The study complied with the Data Privacy Act of 2012 and the National Guidelines for Health and Health-Related Research (2007). The Institution and Ethics Review Board of EAMC reviewed and approved the protocol and all relevant documents before data collection. A letter of authorization was obtained from the Section Head of the Neurology Department prior to access to the medical records. The data were collected and extracted from the patient’s charts from the medical records department; hence, informed consent was not necessary. General data, laboratories, and medications were summarized from the chart and recorded. All the patients’ records were kept and stored in the Medical Records of EAMC in strict confidentiality for at least 6 months. A personal password-protected laptop was used for data collection. The chart information was accessed by the researcher alone and no information was released to a third party. The study did not receive financial grants from agencies, hence, there is no conflict of interest.

## Data Availability

Data cannot be shared publicly because of Data Privacy Act of 2012 and the National Guidelines for Health and Health-Related Research (2007). Data are available from the Medical Records of EAMC for researchers who meet the criteria for access to confidential data.

## Acknowledgments

We thank our mentors and the staff of the Section of Nephrology in the Department of Internal Medicine of the East Avenue Medical Center for their unwavering support.

